# Benefit-Risk Assessment of Medical Products Using Bayesian Multi-Criteria Augmented Decision Analysis for Clinical Development

**DOI:** 10.1101/2023.08.31.23294918

**Authors:** Quang Vuong, Rebecca K Metcalfe, Ofir Harari, Edward J Mills, Jay JH Park

## Abstract

Multi-criteria decision analysis (MCDA) is a benefit-risk assessment tool that evaluates multiple competing benefit and risk endpoints simultaneously. MCDA has the potential to aid sponsors in making effective and informed go/no-go decisions for clinical development programs. MCDA involves assigning weights to benefit and risk endpoints based on their relative importance (i.e., utility weight) and using them to compute a single utility score that represents the overall benefit-risk profile of the treatment. However, to date, MCDA applications have not been appropriate for time-to-event data. In this paper, we introduce a novel framework known as Bayesian Multi-Criteria Augmented Decision Analysis (MCADA) that extends existing probabilistic MCDA methods to encompass time-to-event and ordinal outcomes while incorporating linear and novel non-linear functions in utility aggregation. This paper provides a comprehensive description of the statistical methodology behind the MCADA framework and demonstrates its application using a simulation study, as well as two clinical trials using IPD and aggregate data. Our simulation study found that MCADA generally achieves higher power than the existing MCDA methods due to avoidance of loss of information that occurs when survival and ordinal outcomes are dichotomized. Our two case studies show that the MCADA framework can be effectively used to produce a single utility score that reflects the overall benefit-risk profile of a treatment using both IPD and aggregate data from trials. MCADA broadens the horizon of the current MCDA framework by accommodating a wider range of data types and utility functions in the utility aggregation process.

## 1 Introduction

At the end of an early phase clinical trial, sponsors must decide whether to proceed to the next clinical trial phase (i.e., go/no-go decision) [Chuang-Stein et al., 2011, Glaize et al., 2019, Dharmarajan et al., 2024]. However, optimal go/no-go decision can be impeded by how clinical trials are designed and analyzed. While clinical trials are typically designed around a single primary endpoint, go/no-go decisions require holistic assessments based on multiple outcomes. These decisions are difficult because they require the decision-makers to integrate competing risks and benefits to come to a single decision [Chuang-Stein et al., 2011, Keeney and Raiffa, 1993].

Multi-criteria decision analysis (MCDA) has been proposed as a tool to facilitate these difficult decisions because it can evaluate competing benefit and risk endpoints concurrently [Mussen et al., 2007, Kolasa et al., 2018, Thokala and Duenas, 2012]. Quantitative MCDA integrates benefit and risk endpoints into a single utility score that reflects each endpoint’s relative importance (i.e., utility weight). For example, in linear MCDA each endpoint is multiplied by its utility weight and then the products are summed to calculate a single utility score. This utility score reflects the treatment’s overall benefit-risk profile. The quantitative MCDA framework has the potential to enhance internal decision-making for sponsors during clinical development. Both the European Medicines Agency and the U.S. Food and Drug Administration have supplemented their decision-making processes with MCDA methods, particularly when a multitude of factors need to be considered [Chisholm et al., 2022].

Several MCDA methods have been proposed for probabilistically assessing the benefit-risk profile of medical products [Saint-Hilary et al., 2017, 2018, 2019, Mussen et al., 2008, Tervonen et al., 2011, Nixon et al., 2016, Marcelon et al., 2016]. While these current methods for probabilistic MCDA account for uncertainty in criterion parameters (e.g., Bayesian MCDA [Waddingham et al., 2016]), and accommodate unknown weights or varying preferences (e.g., stochastic multi-criteria acceptability analysis; SMAA [Lahdelma et al., 1998, Tervonen and Lahdelma, 2007, Saint-Hilary et al., 2017]), they have significant limitations. For example, Bayesian MCDA cannot handle ordinal outcomes in their natural form, and neither Bayesian MCDA nor SMAA include appropriate specification for time-to-event outcomes, both of which are common in clinical trials. Using these frameworks as currently applied requires dichotomization of time-to-event outcomes, which is problematic because it can result in loss of information and reduced statistical power to detect a true effect [Altman and Royston, 2006]. Given the prevalence of time-to-event outcomes in clinical trials, there is a need to expand MCDA methods to capture benefit-risk profiles that reflect a broader range of clinical outcomes.

Furthermore, current MCDA approaches often assume linearity and apply a linear utility function when aggregating utilities. This assumption may be unrealistic; for example, assuming linearity for an adverse event would imply that the utility associated with the adverse event decreases constantly which may not be true. Some non-linear utility functions have been proposed [Saint-Hilary et al., 2017, 2018, Menzies et al., 2022], but suffer from unclear clinical interpretation or lack of flexibility. Together, these limitations narrow the versatility of existing MCDA methods and may restrict their use in clinical development decisions.

The aim of this work is to extend existing MCDA approaches to improve the use of outcome information, and consequently statistical power to detect differences between treatments, and clinical interpretation by allowing specification of the utility function. This paper describes a new MCDA statistical approach, Bayesian Multi-Criteria Augmented Decision Analysis (MCADA), that extends existing probabilistic MCDA methods to handle time-to-event and ordinal criteria in their natural forms and allow flexible non-linear utility functions.

To demonstrate MCADA’s capabilities and benefits compared to extant methods we conduct a simulation study and two application studies using data from two oncology clinical trials. These three studies compare MCADA to Bayesian MCDA and SMAA. The paper is organized as follows: Section 2 provides an overview of Bayesian MCDA and SMAA (Table 1). Section 3 introduces MCADA, its technical details, and contributions to MCDA literature. Section 4 discusses the methodological approaches used in our simulation study in accordance with the ADEMP (Aims, Data generating mechanism, Estimands, Methods, and Performance metrics) framework for pre-specification of simulation studies [Morris et al., 2019, Sauer et al., 2024] as well as the details of our case study analyses. Section 5 presents our simulation results comparing MCADA with probabilistic MCDA and SMAA and our case study results applying MCADA to a lung cancer and a pancreatic cancer clinical trial. The paper concludes with a discussion.

**Table 1:** Comparison of MCDA Methods.

| Method | Description | Accepted Outcome Type | Statistical Method of Analysis |
| --- | --- | --- | --- |
| **Bayesian MCDA** [Saint-Hilary et al., 2017]; [Saint-Hilary et al., 2018]; [Waddingham et al., 2016] | Derive utility weights for each criterion based on relative importance. Compute posterior distributions for each criterion, then normalize the distributions. Obtain the final utility score distributions by combining weights with the normalized posterior distributions. | Binary and continuous | Bayesian |
| **SMAA** [Saint-Hilary et al., 2017] | Treat the weights as random variables and draw them from a constrained Dirichlet distribution. Proceed as with the MCDA method. | Binary and continuous | Bayesian |
| **MCADA** | Proceed as with the MCDA method, with additional methodology for time-to-event and ordinal outcomes. | Binary, continuous, time-to-event, and ordinal | Bayesian |

## 2 Overview of Existing Probabilistic MCDA Methods

Probabilistic MCDA is a quantitative benefit-risk assessment tool that help decision-makers evaluate alternatives based on multiple parameters simultaneously while accounting for uncertainty. These methods yield a benefit-risk distribution providing decision-makers with an estimate of the range of probable outcomes (i.e., 95% credible intervals or confidence intervals). Below we describe two probabilistic MCDA methods that account for uncertainty in different analytic inputs: Bayesian MCDA which accommodates uncertainty in the treatment effect; and SMAA which accommodates uncertainty in the treatment effect and the utility weights [Saint-Hilary et al., 2017]. Table 1 summarizes Bayesian MCDA and SMAA, as well as the proposed MCADA method.

### 2.1 Bayesian MCDA

Bayesian MCDA was proposed by Waddingham et al. [2016] and consists of the following steps:

1. Identification of all relevant benefit and risk parameters.
2. Ranking of the different endpoints based on clinical importance to clinicians and patients.
3. Assigning weights to each endpoint based on the ranking from step 2.
4. Deriving posterior distributions for each endpoint using Bayesian analysis, in which parameters are treated as random variables and conditioned on observed data.
5. Normalizing each vector of posterior samples derived in Step 4 to values between 0 and 1.
6. Calculating utility scores distributions by combining the weights and normalized distributions.

### 2.2 Stochastic Multi-Criteria Acceptability Analysis (SMAA)

Stochastic multi-criteria acceptability analysis (SMAA), first proposed by Lahdelma et al. [1998], is an extension of probabilistic MCDA. SMAA is useful when no information is provided on the relative importance of each decision endpoint, or when there is uncertainty about each endpoint’s importance.

Like probabilistic MCDA, SMAA is limited to continuous and binary criteria. Instead of inputting a fixed vector of weights *w*_*j*_ as random variables. The space of weights *W* is defined as an *n* − 1-dimensional simplex in *n*-dimensional space:

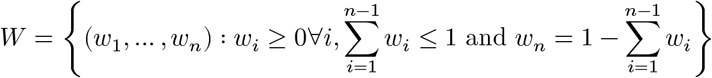

The weights are drawn from a Dirichlet distribution and constrained within a simplex such that the sum of the weights is equal to 1. Then, the utility scores are random variables, and their distributions can be obtained by sampling values from the distributions of the endpoint parameters and uniformly from the space of weights *w*. Under the SMAA model, the utility score distributions account for both sampling variation in the endpoint measurements and uncertainty in the preferences of the endpoints. However, the benefit-risk profiles of treatments can only be compared when the same weight vector is used; profiles cannot be compared over the entire weight space. Thus, the distribution of the difference between two utility scores can be computed by simulating values for the weights in the weight space. Note that *W* may be constrained to incorporate preferences from decision makers.

## 3 Overview of MCADA

In this section, we provide the technical details of MCADA and discuss how MCADA extends the MCDA methodology to allow the evaluation of outcomes beyond binary and continuous endpoints, and to allow the modelling of non-linear utility functions. MCADA is a direct extension of Bayesian MCDA proposed by Waddingham et al. [2016].

For binary, continuous and ordinal variables, given *m* treatments (*i* = 1, …, *m*) assessed via *n* different endpoints (*j* = 1, …, *n*), the performance of treatment *i* on endpoint *j* is denoted by *ξ*_*ij*_. Probabilistic MCDA, SMAA, and MCADA treat *ξ*_*ij*_ as a random variable and uses Bayesian modelling to assign a probability distribution to each *ξ*_*ij*_, depending on the data type of the endpoint. If endpoint *j* produces binary data, *ξ*_*ij*_ will be assumed to follow a beta distribution arising from a beta-binomial model with parameters determined by the number of events and patients per treatment, *ξ*_*ij*_ ~ *Beat*(*α*_*ij*_, *β*_*ij*_). If it is continuous, *ξ*_*ij*_ will be assumed to follow a Student-t distribution informed by the mean, standard deviation, and number of patients assessed by endpoint *j*. When using MCADA, ordinal and time-to-event endpoints may also be modelled. If *j* is an ordinal variable, *ξ*_*ij*_ will be assumed to follow a Dirichlet distribution arising from a Dirichlet-multinomial model. The Dirichlet parameters will be informed by both the prior and the data, where the data are summarized as a vector of counts per category. For each treatment *i*, we have a vector indicating how the treatment performed on each criterion: *ξ*_*i*_ = (*ξ*_*i*1_, …, *ξ*_*in*_).

If endpoint *j* is a time-to-event variable, using MCADA, the posterior of *ξ*_*ij*_ will be computed by first fitting a parametric survival curve based on a dependent gamma prior, then by calculating the median survival time for each posterior draw of individual survival curves.

To compute the posterior survival function for time-to-event data, we use the approach of Castillo and Van der Pas [Castillo and van der Pas, 2021]. This method uses a dependent gamma prior to inform the hazard rates of a piecewise exponential prior. Given the time interval of the data, [0, *τ*], divide the interval into *K* intervals of equal size: *I*_*k*_, *k* = 1, …, *K*. The value of the hazard function *λ* over each sub-interval is assigned a piecewise constant Gamma(*α*, *β*) prior, with shape parameter *α* and rate parameter *β*. The prior on the hazard *λ* is

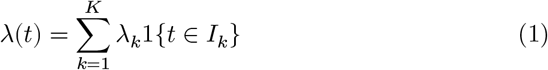

With

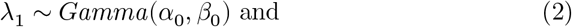

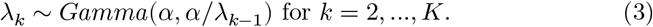

The dependent structure of the gamma prior can be seen from the mean and variance for *k* = 2, …, *K*:

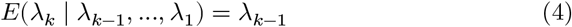

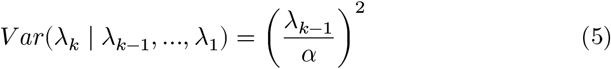

The equally sized intervals allow the theoretical properties of the posterior distribution of the survival curves to be derived, but it is possible to use differently sized intervals if desired. The piecewise and dependent structure of the hazard function introduces some smoothness to the hazard function while modelling the survival times non-parametrically.

### 3.1 Partial Value Function

Once posterior samples have been drawn for each *ξ*_*ij*_, the sampled posterior values are passed through monotonically increasing partial value functions, *u*_*j*_(·), that maps them onto the [0, 1] interval. That is, if 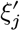 denotes the most desirable value for endpoint *j* and 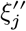 the least desirable value, we have 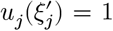 and 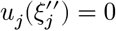.All other utility values are given by

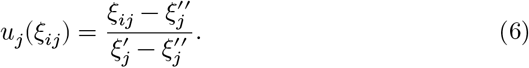

Here, *u*_*j*_(*ξ*_*ij*_) > *u*_*j*_(*ξ*_*hj*_) indicates that treatment *i* is preferred to treatment *h* for endpoint *j*.

This linear partial value function, while a common choice in decision analysis, has been shown in some cases to recommend a treatment with either extremely high risk or low benefit [Menzies et al., 2022]. To address these issues, we introduced two non-linear transformations of the partial value functions: the Emax function and the logistic function. The Emax function is a non-linear function used for estimating dose-response curves [Macdougall, 2006]. In the context of MCADA, instead of measuring a dose-response relationship, we use the monotone, concave shape of Emax to measure a performance-utility relationship. Emax requires two inputs, *U*_0_ and *U*_50_, where *U*_0_ is the expected utility when the performance is lowest and *U*_50_ is the performance that produces half of the maximum utility value. These parameters can be adjusted based on the clinical expectation of the performance-utility relationship in settings where this association is non-linear. The logistic function requires the same parameter inputs as Emax, with the addition of parameter δ, that determines the steepness of the logistic slope. The Emax function transformation of the partial value function is

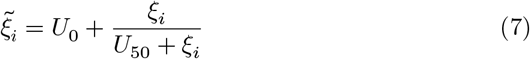

and the logistic function is

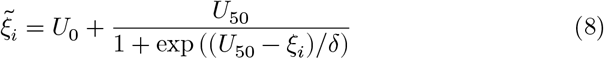

The linear and non-linear partial value functions used in MCADA can be seen in Figure 1.

**Figure 1.**
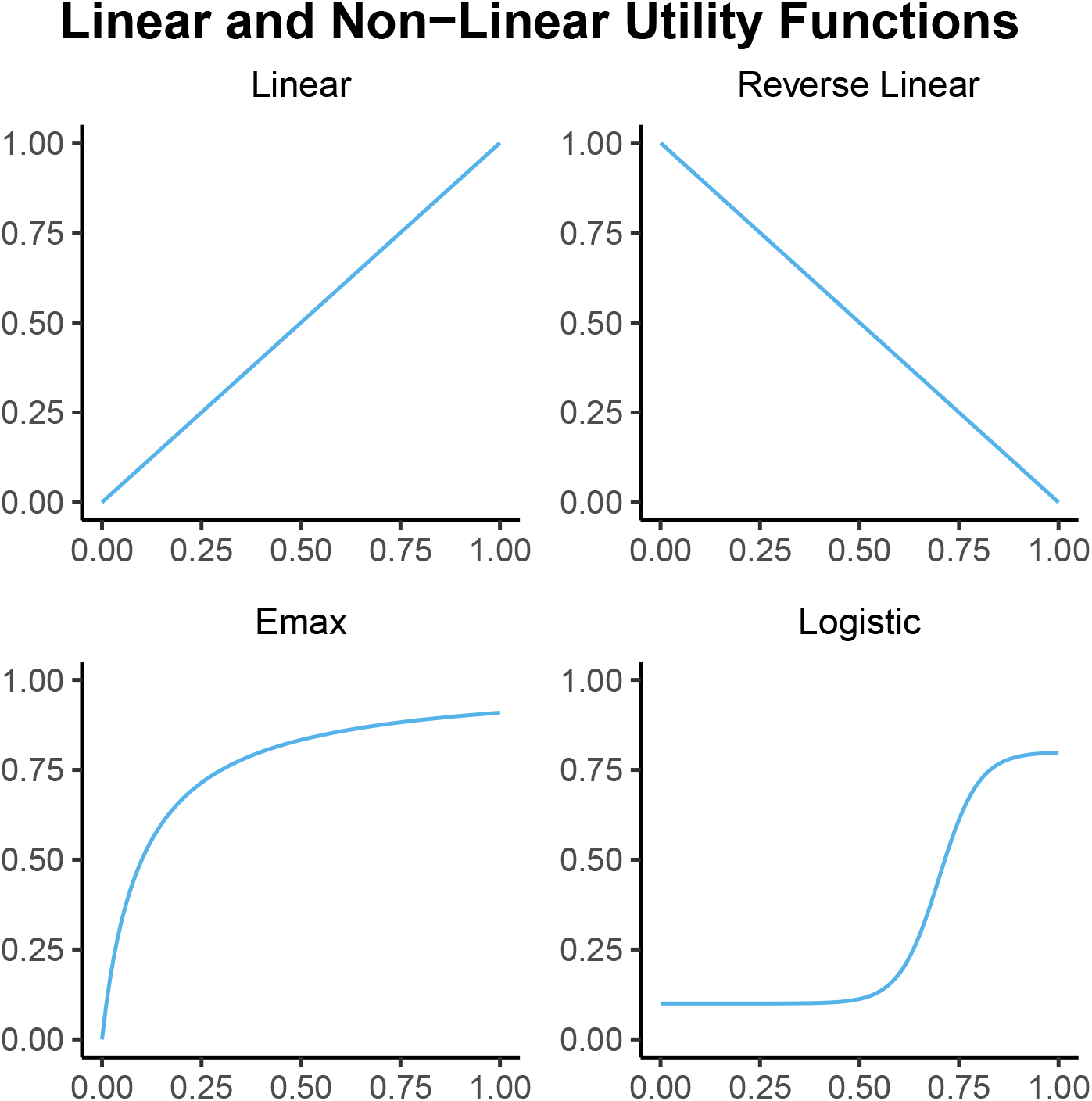
Example linear and non-linear partial value function shapes. Parameter values to generate the Emax curve: *U*_0_ = 0 and *U*_50_ = 0.1. Parameter values to generate this logistic curve: *U*_0_ = 0.1, *U*_50_ = 0.7, and δ = 0.05.

### 3.2 Utility Aggregation

Using the vector *w* = (*w*_1_, …, *w*_*n*_) provided by the stakeholder to assign weights to the respective criteria, a linear utility score *u*(*ξ*_*i*_, *w*) is then calculated for treatment *i* as a weighted average

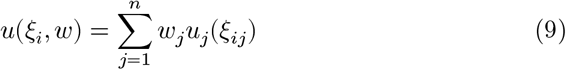

Under the Bayesian model, the utility score is in fact a random variable, whose samples are obtained from repeated sampling from the posteriors of the *ξ*_*ij*_. For non-linear transformations, simply replace *ξ*_*i*_ with 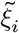 in the above equation.

The utility scores for different treatments are compared graphically and through descriptive statistics.

### 3.3 Treatment Recommendation

To generate a treatment recommendation, treatments are compared pairwise using the MCADA approach. The utility score of Δ*u* (*ξ*_*i*_, *ξ*_*h*_, *w*), the utility difference between treatments *i* and *h*, can be taken advantage of. Let *x*_*i*_ = (*x*_*i*1_, … *x*_*in*_) and *x*_*h*_ = (*x*_*h*1_, … *x*_*hn*_) denote observed outcomes, where each element of *x*_*i*_ and *x*_*h*_ corresponds to either a vector of patient-level data or a scalar of aggregate-level data for treatment *i* and *h* respectively, depending on what data is available for analysis. Then, given observed outcomes *x*_*i*_ = (*x*_*i*1_, … *x*_*in*_) and *x*_*h*_ = (*x*_*h*1_, … *x*_*hn*_) with corresponding treatment performances *ξ*_*i*_ and *ξ*_*h*_, recommendation with regard to the preferability of treatment *i* over treatment *h* will rely on

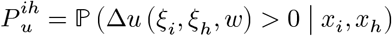

that is: the posterior probability of utility gain, where the subscript *u* in 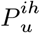 refers to the use of the utility score from the previous equation. Note that 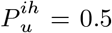 corresponds to treatments *i* and *h* having equal benefit-risk ratios. A natural extension to make a decision based on this probability, suggested by [Menzies et al., 2022], would be to compare it to a threshold confidence level 0.5 < *ψ* ≤ 1. Then, 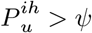 indicates that there is enough evidence to support *i* as having a better benefit-risk ratio than *h* with confidence level *ψ*.

## 4 Methods

In the applications and the simulation described below, we considered three common endpoints used in oncology clinical trials, specifically survival, measures of tumor burden, and incidence of adverse events. These were operationalized as:

- Overall survival (OS), a time to event variable
- Response evaluation criteria in solid tumors (RECIST), [Eisenhauer et al., 2009] an ordinal outcome
- Presence of grade 3 or 4 adverse events, a binary outcome

### 4.1 Simulation Study

Our simulation study was conducted using the following steps: 1. Simulate randomized clinical trials with two treatments, *T*_1_, *T*_2_, each with three uncorrelated endpoints of sample size *N* = 200. Treatment *T*_1_ is considered the treatment, while treatment *T*_2_ is considered the control. The three endpoints are: one binary safety endpoint; one ordinal efficacy endpoint; and one time-to-event efficacy endpoint. 2. Derive the posterior distributions of the endpoint parameters using the simulated data assuming a flat prior. Draw 10,000 samples from each posterior distribution and obtain the empirical distributions by passing the values through the partial value function. 3. Use the partial value function output to compute the probability that treatment *T*_1_ has a better benefit-risk profile 𝒫^1,2^ than treatment *T*_2_. If 𝒫^1,2^ > *ψ*, then treatment *T*_1_ is recommended. If 𝒫^1,2^ < 1 − *ψ*, then treatment *T*_2_ is recommended. If 1 − *ψ* ≤ 𝒫^1,2^ ≤ *ψ*, then neither treatment is recommended. 4. Repeat steps 1-3 for 2,500 simulated trials. 5. Estimate the probability that each treatment is recommended ℙ(𝒫^1,2^ > *ψ*) by its proportion over 2,500 simulated trials.

The Bayesian MCDA and SMAA methods follow the above steps, with an additional dichotomization step between the first and second steps. The ordinal RECIST outcome was converted to an objective response rate (yes/no) by summing the rates of complete response (CR) and partial response (PR) as having responded to the treatment, and stable disease (SD) and progression (PD) as not responding to the treatment. For OS, we considered a landmark analysis at 12 months using the same simulated time-to-event data that was used for MCADA.

Further details of the simulation study are described below in accordance with the ADEMP framework [Morris et al., 2019].

#### 4.1.1 Aims

The goal of this simulation study was to compare the performance measured by the statistical power of Bayesian MCDA, SMAA, and MCADA based on common criteria used in oncology clinical trials.

#### 4.1.2 Data Generating Mechanism

All the parameters for generating OS, RECIST, and adverse event variables are described in Tables 2 and 3. In particular, Table 2 specifies the simulation parameters for each criterion in the treatment and control groups under the null hypothesis (“equal”) and alternative hypothesis (“treatment superior”), while Table 3 specifies which criteria that are equal or superior in the treatment in the simulated scenarios. The time-to-event OS data was generated using an exponential distribution with rate *λ* specified for the treatment and the control groups under the null and alternative scenarios. A random time to censoring variable was generated according to a log-normal distribution with mean and standard deviation on the log scale. Observations for which the time-to-event was greater than the censoring time were considered as right-censored. The true hazard rate (HR) was set to 1 for controls and to 0.9 for the treated. The ordinal variable RECIST was generated using a multinomial distribution with probabilities π_1_, π_2_, π_3_, and π_4_ specified for the controls and treated under the null and alternative scenarios. The cell probabilities were chosen to correspond to an odds ratio (OR) of 0.85 in a proportional odds logistic model under the alternative scenario. We also assumed a vector of weights 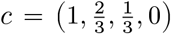 for complete response, partial response, stable disease and progression, respectively, to reflect the relative desirability of the different outcomes. The adverse events binary variable was generated using a binomial distribution with a probability of success π specified for controls and the treated under the null and alternative scenarios. The RR was set to 1 for the controls and to 0.9 for the treated.

**Table 2:** Assumptions.

| Scenario | Study Arm | Control | Treatment | Treatment effect |
| --- | --- | --- | --- | --- |
| Grade 3 and 4 Adverse Event Rate | Null Scenario | 0.4 | 0.4 | RR: 1.00 |
|  | Alternative Scenario | 0.4 | 0.36 | RR: 0.90 |
| RECIST Response Rate (CR, PR, SD, PD) | Null Scenario | (0.102, 0.7, 0.102, 0.096) | (0.102, 0.7, 0.102, 0.096) | OR: 1.00 |
|  | Alternative Scenario | (0.102, 0.7, 0.102, 0.096) | (0.118, 0.687, 0.088, 0.107) | OR: 0.85 |
| Overall Survival (Median OS) | Null Scenario | 12 | 12 | HR: 1.00 |
|  | Alternative Scenario | 12 | 13.33 | HR: 0.90 |

**Table 3:** Simulation Scenarios.

| Scenarios | AE | RECIST Criterion | Overall Survival |
| --- | --- | --- | --- |
| 1 | Equal | Equal | Equal |
| 2 | Treatment superior | Equal | Equal |
| 3 | Equal | Treatment superior | Equal |
| 4 | Equal | Equal | Treatment superior |
| 5 | Treatment superior | Treatment superior | Equal |
| 6 | Treatment superior | Equal | Treatment superior |
| 7 | Equal | Treatment superior | Treatment superior |
| 8 | Treatment superior | Treatment superior | Treatment superior |

#### 4.1.3 Estimand

The primary quantity of interest (estimand) was the difference in mean utility score between the treatment and control arm. This estimand for each arm was calculated based on OS, RECIST, and adverse event parameters that were weighted by their relative importance. Note when interpreting the term “estimand” that this study follows the ADEMP framework [Morris et al., 2019, Sauer et al., 2024].

#### 4.1.4 Methods

We compared the Bayesian MCDA, SMAA, and MCADA methods in this simulation study. The weighting scheme of grade 3 or higher adverse event rates (binary), RECIST criterion (ordinal), and OS (time-to-event) for MCADA and Bayesian MCDA method was derived using analytical hierarchy process (AHP) [Saaty, 1987]. Treatment performance on OS was assumed to be twice as important as the RECIST criteria, and the RECIST was considered twice as important as adverse event rates. Weights of 0.57, 0.29, and 0.14 were assumed for OS, RECIST criterion (tumor response), and grade 3 or 4 adverse events, respectively. The weights for each endpoint are summarized in Table 4. As a sensitivity analysis, we also considered a scenario where all criteria were assumed to be of equal importance and assigned a weight of 0.33.

**Table 4:** Summary of variables and weights used in simulations.

|  | Overall Survival |  | RECIST Tumor Criterion |  | Grade 3 or 4 Adverse Events |  |
| --- | --- | --- | --- | --- | --- | --- |
|  | Definition | Weights | Definition | Weights | Definition | Weights |
| MCADA | Time to event: OS | 0.57 | Ordinal, 4 levels: complete response (CR); partial response (PR); stable disease (SD); progressive disease (PD) | 0.29 | Binary: presence or absence of at least one grade 3 or 4 adverse event | 0.14 |
| MCDA | Binary: death observed at 12 months (yes/no) | 0.57 | Binary: Objective response rate | 0.29 | Binary: presence or absence of at least one grade 3 or 4 adverse event | 0.14 |
| SMAA | Binary: death observed at 12 months (yes/no) | Simplex Sampling<sup>†</sup> | Binary: Objective response rate | Simplex Sampling | Binary: presence or absence of at least one grade 3 or 4 adverse event | Simplex Sampling |

We designed our simulation study based on patient-level data of an early phase randomized clinical trial on extensive-disease small cell lung cancer (NCT01439568) that collected data on OS, RECIST, and adverse events.

We considered two scenarios with differing strategies for false positive error rate control. As the base case scenario, we calibrated each MCDA method to have a type I error rate controlled at 0.10. For each method, we calibrated the superiority threshold *ψ* defined by the posterior probability of the difference in the utility score between the treatment and control being greater than 0.10. This was done by simulating data under the null scenario where the treatment was assumed to have no effect across all three parameters of OS, RECIST, and adverse events.

As the supplementary scenario, we considered the same superiority thresholds of 0.95 across the three MCDA methods.

#### 4.1.5 Performance Measures

The main performance metric for our simulation study was the probability of recommending the treatment over the control. This is interpreted as the type I error rate in the null hypothesis scenario and power (1 - type II error rate) in alternative hypothesis scenarios. We considered seven alternative hypothesis scenarios, in which treatment was assumed to be superior to the control on at least one parameter (Table 3). We considered that treatment could reduce the relative risk (RR) for adverse event (binary outcome) by 10% (RR: 0.90). The treatment effects of 0.85 odds ratio (OR) and 0.90 hazard ratio (HR) were assumed for RECIST (ordinal) and OS (time-to-event) parameters, respectively. For each scenario, we simulated 2,500 trials, and for each trial, 10,000 posterior samples were drawn to obtain the treatment recommendation.

### 4.2 Case Studies

In this section, the use of MCADA to support decision-making for clinical development programs in oncology is illustrated using aggregate data and IPD from two early phase trials obtained from Project Data Sphere [Green et al., 2015].

The first trial was an open-label, randomized early phase}= trial conducted to evaluate the clinical efficacy and safety of a novel selective peptide antagonist, LY2510924, in combination with standard of care (SOC; i.e. carboplatin/etoposide) compared to SOC alone in patients with extensive-disease small cell lung cancer was (NCT01439568) [Salgia et al., 2017]. A total of 94 patients were randomized to receive the novel therapy in combination with standard-of-care (N=47) or standard-of-care alone (N=43). Full details of the event rates by arm are shown in Table 5.

**Table 5:** Summary of statistics from extensive-disease small cell lung cancer trial.

| Statistics | Treatment (n=47) | Control (n=43) |
| --- | --- | --- |
| Overall Survival, median (95% CI) (in months) | 9.72 (6.6, 11.7) | 11.1 (8.3, 13.4) |
| RECIST Response, n (%) | 35 (86) | 34 (81) |
| Grade 3 or 4 Adverse Event, n (%) | 24 (51) | 13 (30) |

Abbreviation: confidence interval, CI.

The second trial was a randomized, multicenter early phase trial conducted to evaluate the safety and efficacy of CO-101, a lipid-drug conjugate (intervention), compared to gemcitabine (control) among patients with untreated metastatic pancreatic ductal adenocarcinoma and lower hENT1 expression (NCT01124786). A total of 367 patients were randomized at an equal ratio (1:1) to CO-101 (treatment) versus gemcitabine (control). This trial was powered to detect differences in OS. The details of the event rates by arm are shown in Table A1 of the Supplementary Materials.

Two benefit outcomes of IPD data, OS and RECIST criteria, as well as one aggregate data risk outcome, grade 3 or higher adverse event rates, were considered for both case studies. Utility weights were derived using AHP [Saaty, 1987] and were the same as in the simulation study: weights of 0.57, 0.29, and 0.14 were assumed for OS, RECIST criteria, and grade 3/4 adverse events, respectively. As a sensitivity analysis, all outcomes were assumed to be of equal importance and were assigned a utility value of 0.33.

For OS, the most desirable median survival time was set at 20 months, and the least desirable median survival time was set at 0 months. For RECIST criteria, the best response recorded was considered, and relative utility values of 0, 0.33, 0.67, and 1 were assigned to progressive disease, stable disease, partial response, and complete response, respectively. For the adverse event rate, the most desirable value was 0% and the least desirable value was 50% or higher, so a utility value of 0 would be assigned if at least 50% of trial participants experience an adverse event.

For both case studies, we present utility distributions derived using the linear utility function. The non-linear performance-utility relationship for OS and adverse event rates, as well as the linear relationship for RECIST response is visualized in Figure 2. For all non-linear results, the Emax function with parameters *U*_0_ = 0 and *U*_50_ = 0.3 was selected for OS, and the logistic function with parameters *U*_0_ = 0.25, *U*_50_ = 0.5, and δ = 0.05 was selected for adverse event rates. The resulting utility distributions from the non-linear functions can be found in the Supplementary Materials.

**Figure 2.**
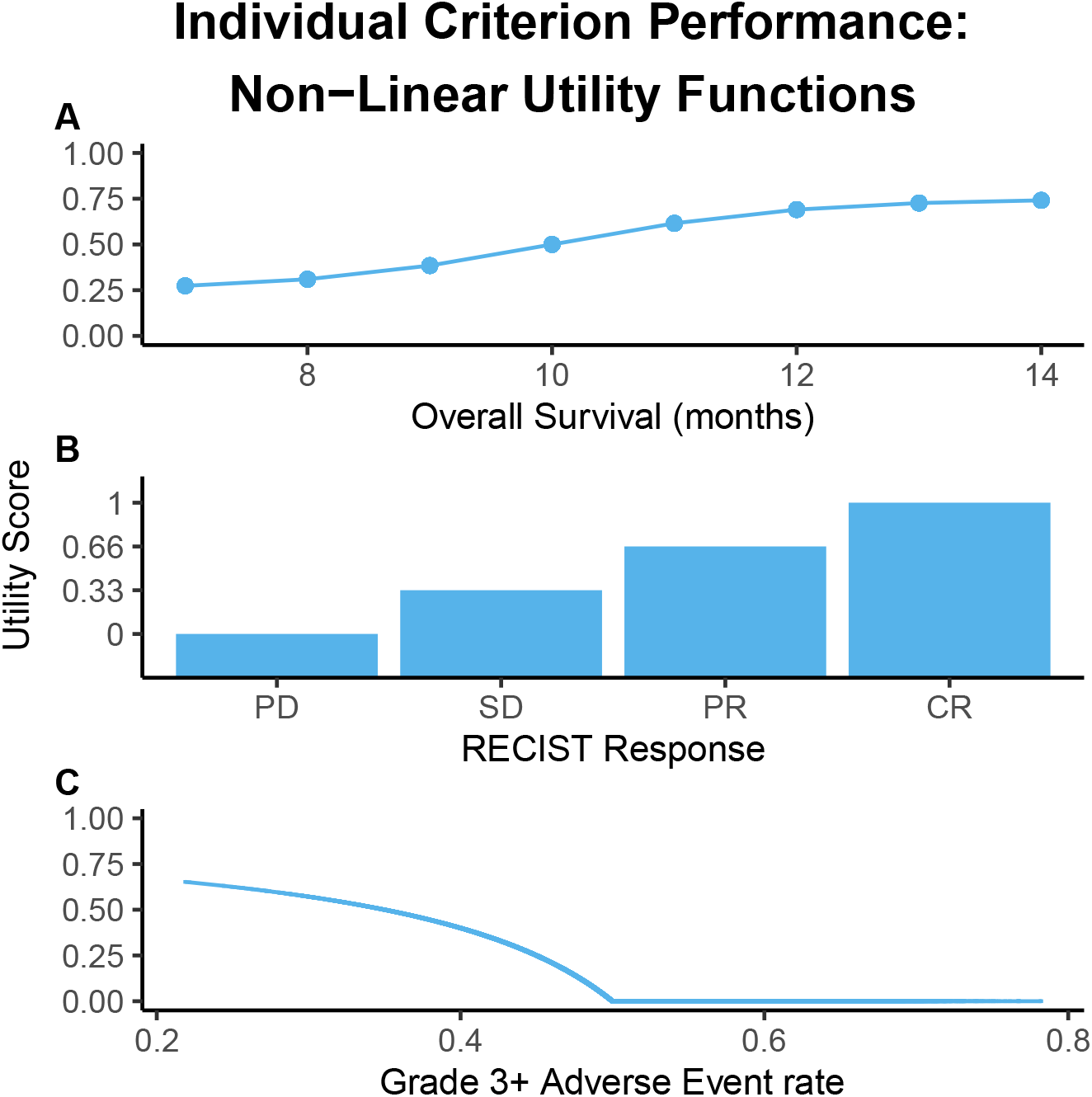
The performance-utility relationship for A) overall survival, B) RECIST response, and C) grade 3+ adverse events. A) results from a logistic transformation with *U*_0_ = 0.25, *U*_50_ = 0.5, and δ = 0.05, B) results from a linear function, and C) results from an Emax transformation with *U*_0_ = 0 and *U*_50_ = 0.3.

### 4.3 Software

We used R version 4.3.2 [R Core Team, 2023] to conduct the simulation studies and data analyses of the case studies. We implemented MCADA and re-programmed Bayesian MCDA and SMAA in R scripts provided in the online Supplementary Materials.

## 5 Results

### 5.1 Simulation Study Results

For the base case scenario, we compared the performance of MCADA to Bayesian MCDA and SMAA as measured by statistical power, where the type I error rate was controlled at 10% (Table 6). As we considered OS to be the most important parameter, alternative hypothesis scenarios where treatment was assumed to affect OS had higher power than the other scenarios. This trend was observed across all three methods.

**Table 6:** Simulation results: type I error rate and power. Monte Carlo standard errors are in brackets.

| Scenario |  | MCADA | MCDA | SMAA |
| --- | --- | --- | --- | --- |
|  |  | Threshold 0.95 | Threshold 0.895 | Threshold 0.84 |
| 1 | Type I error rate | 0.100 (0.006) | 0.100 (0.006) | 0.099 (0.006) |
| 2 | Only safety better for Treatment | 0.146 (0.007) | 0.145 (0.007) | 0.167 (0.007) |
| 3 | Only RECIST better for Treatment | 0.006 (0.002) | 0.001 (0.001) | 0.001 (0.001) |
| 4 | Only OS better for Treatment | 0.910 (0.006) | 0.889 (0.006) | 0.468 (0.010) |
| 5 | Safety and RECIST better for Treatment | 0.143 (0.007) | 0.169 (0.007) | 0.219 (0.008) |
| 6 | Safety and OS better for Treatment | 0.944 (0.005) | 0.916 (0.006) | 0.665 (0.009) |
| 7 | RECIST and OS better for Treatment | 0.921 (0.005) | 0.869 (0.007) | 0.463 (0.010) |
| 8 | Treatment better in all 3 parameters | 0.956 (0.004) | 0.889 (0.006) | 0.704 (0.009) |

In general, MCADA had higher power compared to Bayesian MCDA and SMAA. In scenario 8, where treatment was assumed to be better than the control on all three parameters, MCADA showed the highest statistical power (1 − *β* = 0.956), followed by Bayesian MCDA and SMAA (1 − *β* = 0.889 and 0.704, respectively). Figure 3 shows the control and treatment utility scores using MCADA for scenario 8. The relative utility scores estimated for scenario 8 for each of the three methods are illustrated in Figure 4.

**Figure 3.**
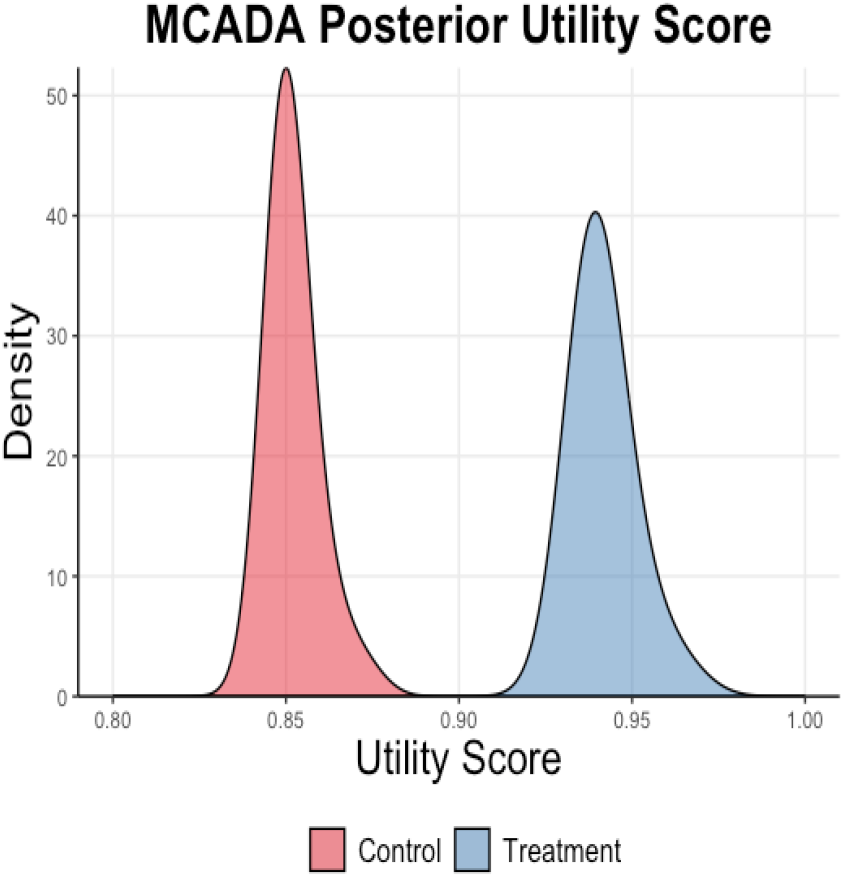
MCADA posterior utility scores under scenario 8: treatment superior to control in all three parameters

**Figure 4.**
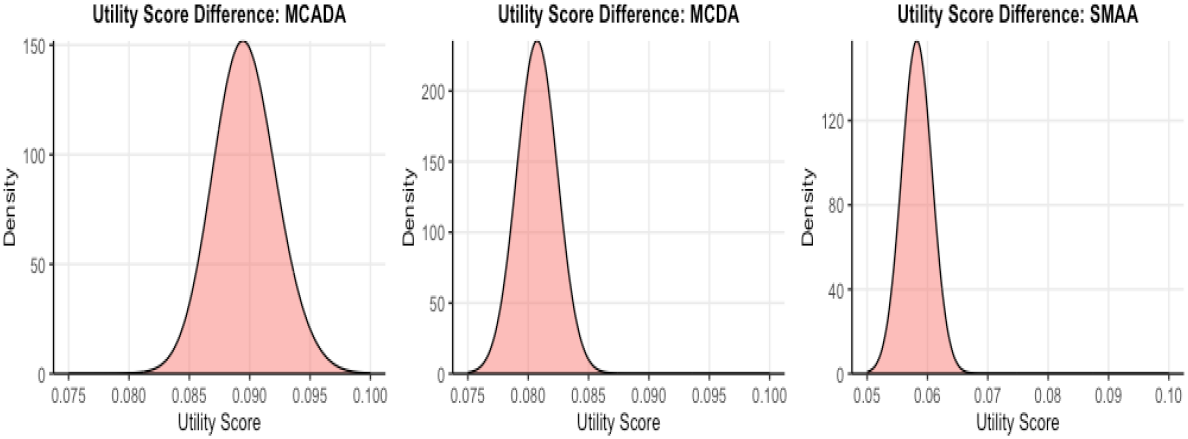
Difference between control and treatment arm utility scores for MCADA, Bayesian MCDA, and SMAA under scenario 8: treatment superior to control in all three parameters.

We conducted two supplementary scenarios in our simulation. We compared the performance of the three MCDA methods using the same posterior superiority threshold of 0.95 with different utility weights (Table A3 in the Supplementary Materials). We performed additional simulations with equal utility weights for OS, RECIST, and adverse events (Table A4 in the Supplementary Materials). Our supplementary cases showed similar findings as the base case. In general, MCADA outperformed Bayesian MCDA and SMAA across different scenarios in terms of statistical power.

### 5.2 Case Study 1: Extensive-Disease Small Cell Lung Cancer

For the first case study, the calculated utility score for the experimental treatment arm was lower (0.47; 95% credible interval [CrI]: 0.41, 0.53) than the control arm (0.56; 95% CrI: 0.47, 0.69) (Table 7). The results of the MCADA analysis showed that the novel therapy added no clinical utility when compared to the standard-of-care (Figure 5). Similarly, when using the non-linear functions for OS and adverse events, the mean utility score for the treatment arm was lower (0.47; 95% CrI: 0.39, 0.58) than the control arm (0.60; 95% CrI: 0.44, 0.69) (Figure A2 in Supplementary Materials).

**Table 7:**
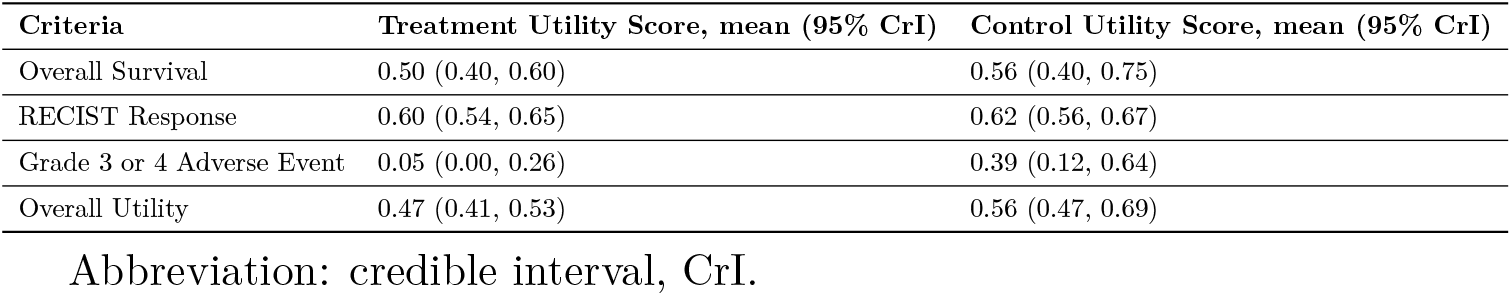
Summary of utility score results from case study 1 using linear utility functions.

| Criteria | Treatment Utility Score, mean (95% CrI) | Control Utility Score, mean (95% CrI) |
| --- | --- | --- |
| Overall Survival | 0.50 (0.40, 0.60) | 0.56 (0.40, 0.75) |
| RECIST Response | 0.60 (0.54, 0.65) | 0.62 (0.56, 0.67) |
| Grade 3 or 4 Adverse Event | 0.05 (0.00, 0.26) | 0.39 (0.12, 0.64) |
| Overall Utility | 0.47 (0.41, 0.53) | 0.56 (0.47, 0.69) |

Abbreviation: credible interval, CrI.

**Figure 5.**
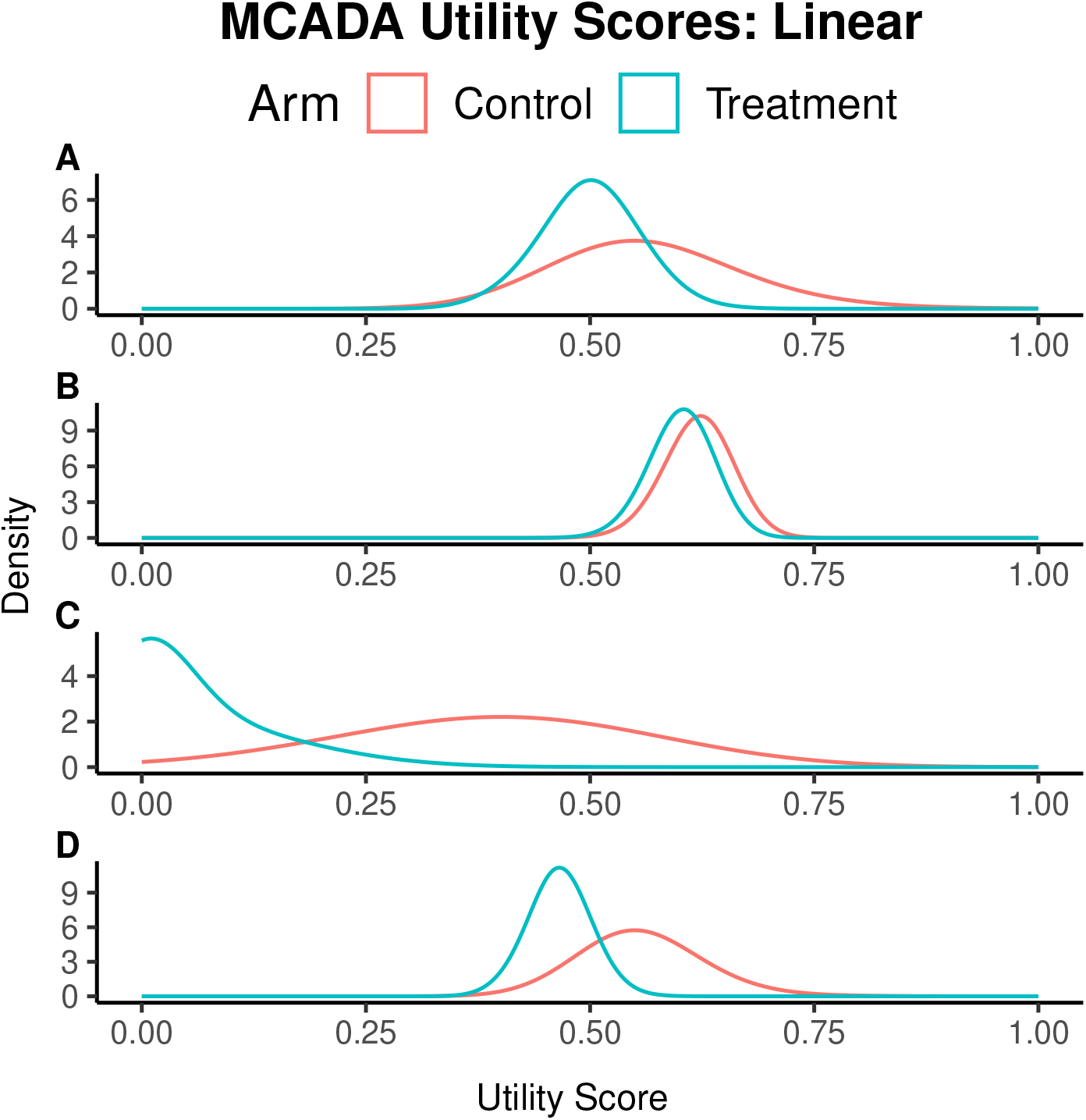
The utility score distributions of LY+SOC (treatment) and SOC (control) for (A) overall survival, (B) best tumor response measured by RECIST, (C) grade 3 or 4 adverse events, and (D) overall utility in patients with extensive disease non-small cell lung cancer. The utility values (relative importance) of 0.57, 0.29, and 0.14 were assumed for overall survival, best tumor response measured by RECIST, and grade 3 or 4 adverse event, respectively.

For the intervention, the mean (95% CrI) utility score was 0.50 (0.40 0.60) for OS, 0.65 (0.62, 0.68) for tumor response, and 0.05 (0.00, 0.26) for adverse events.

For SOC, the mean (95% CrI) was 0.56 (0.40, 0.75) for OS, 0.62 (0.56, 0.67) for tumor response, and 0.39 (0.12, 0.64) for adverse events (Figure 5). When computing the utility scores using non-linear functions, the Emax function was used for OS and the logistic function was used for adverse events. For the intervention arm, the mean (95% CrI) utility score was 0.50 (0.31, 0.69) for OS and 0.10 (0.00, 0.46) for adverse events. For the SOC arm, the mean (95% CrI) was 0.60 (0.44, 0.69) for OS and 0.54 (0.28, 0.68) for adverse events. Comparing the utility score between intervention and control arms of each component independently using both linear and non-linear functions reiterates that SOC had a superior benefit-risk profile compared to the treatment (Figure 5).

The difference in utility scores between treatment and SOC was computed: the mean (95% CrI) difference in utility was −0.09 (−0.22, 0.02). The utility score distributions were also computed using equal weighting for each outcome, as a sensitivity analysis (see Supplementary Materials). Results of the sensitivity analysis showed that the mean utility score (95% CrI) was 0.38 (0.34, 0.49) for the intervention arm and 0.52 (0.41, 0.63) for the SOC arm reiterating SOC has a superior benefit-risk profile than the intervention.

### 5.3 Case Study 2: Metastatic Pancreatic Ductal Adenocarcinoma

For the second case study, the median OS was shorter for the intervention arm compared to the control arm but the difference was not statistically different (hazard ratio [HR] [95% CI]: 1.072 (0.86, 1.34); median [95% CI] 5.2 (4.5, 6.2) months for treatment arm and 6.0 (5.2, 6.8) months for control arm).

Overall response rate was 16.2% (N=25/154) for the intervention arm and 28.8% (N=41/142) for the control arm. The grade 3 or 4 adverse event occurred in 45.8% (N=82/179) of participants in the intervention arm, and 45.3% (N=82/181) in the control arm.

The overall utility score that accounts for OS, tumor response, and adverse events for CO-101 versus gemcitabine is shown in Figure 6. Although the primary outcome of OS alone did not provide a clear indication of whether the treatment or control arm was superior, the differences between the benefit-risk profile of the intervention and the control arm became more evident when examining the MCADA results. Using MCADA found that the intervention had a much lower mean overall utility score[95% CrI] (0.44 (0.38, 0.50)) than the gemcitabine (control) arm (0.50 (0.43, 0.58)), indicating that the intervention did not show any additional clinical utility compared to control (Figure 6; Table A2).

**Figure 6.**
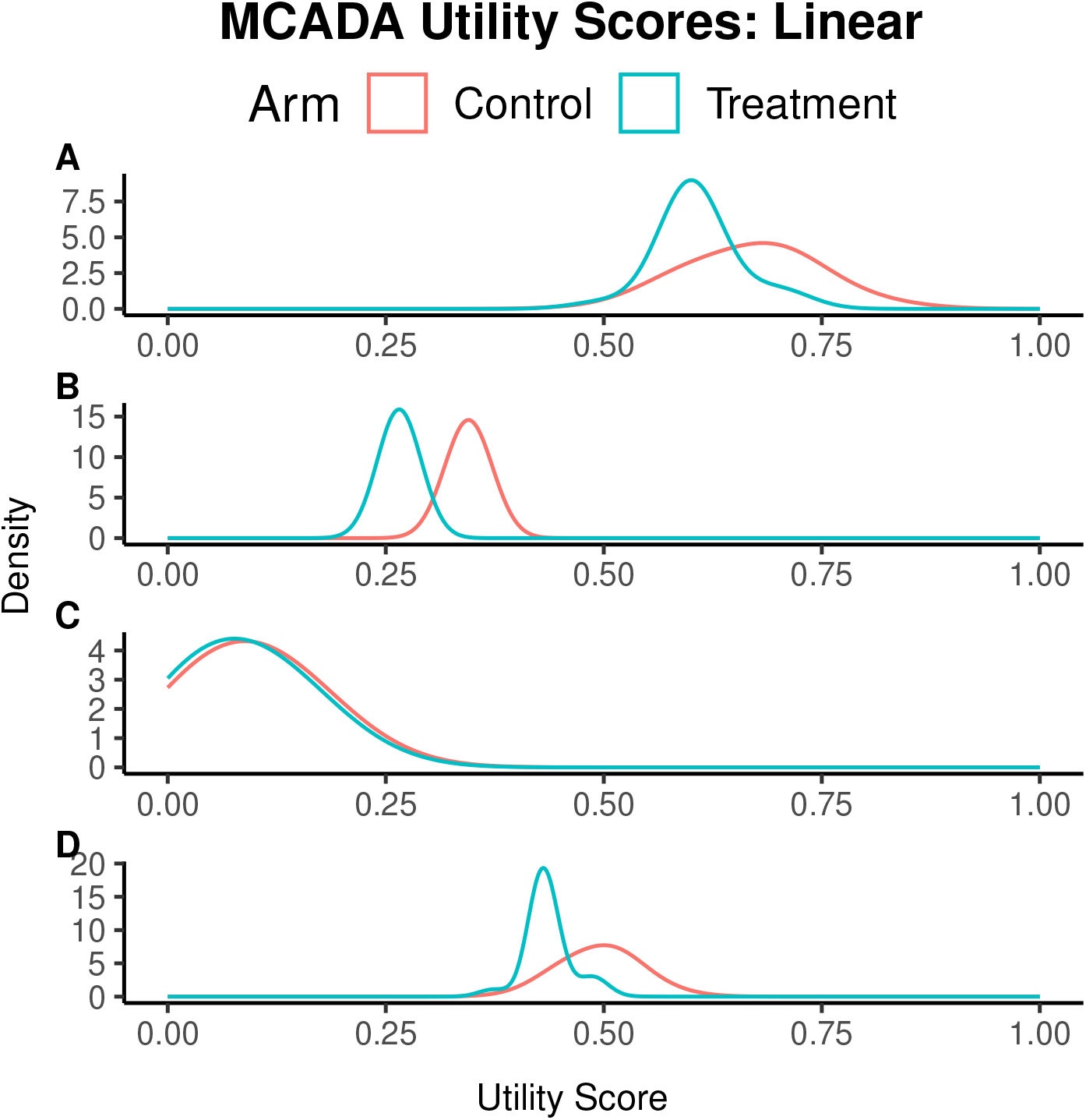
The utility score distributions of CO-101 (treatment) and gemcitabine (control) for (A) overall survival, (B) best tumor response measured by RE-CIST, (C) grade 3 or 4 adverse events, and (D) overall utility in patients with metastatic pancreatic ductal adenocarcinoma. Utility values (relative importance) of 0.57, 0.29, and 0.14 were assumed for overall survival, RECIST criteria, and grade 3 or 4 adverse events, respectively.

Comparing the individual criterion performances again demonstrated the superiority of the control arm compared to the treatment arm. For the intervention, the mean (95% CrI) utility score was 0.61 (0.50, 0.70) for OS, 0.27 (0.23, 0.30) for RECIST response, and 0.09 (0.00, 0.23) for adverse event. For the control, the mean (95% CrI) was 0.67 (0.60, 0.80) for OS, 0.34 (0.31, 0.38) for RECIST response, and 0.10 (0.00, 0.24) for adverse event (Figure 6).

The difference in utility scores between the treatment and control arms was also computed, and the mean (95% CrI) difference in utility was −0.06 (−0.15, 0.04). All utility score computations showed that the control had a superior benefit-risk profile compared to the intervention.

## 6 Discussion

Clinical trial decisions are often based on a single primary outcome, making go/no-go decisions challenging when results are inconclusive. To help facilitate decisions based on a more comprehensive evaluation of treatment performance, MCDA approaches are often used. This paper introduces the MCADA method that can handle time-to-event and ordinal outcomes and non-linear utility functions for aggregation. We demonstrate the practical application of MCADA using a simulation study and analyses of two clinical trial datasets in oncology.

Using simulated data, we compare the statistical power of MCADA to Bayesian MCDA and SMAA and demonstrate that MCADA improves statistical power. Consequently, MCADA is better able to detect differences in risk-benefit profiles of medical products, as measured by utility scores. These findings are not surprising as MCADA retains the natural form of time-to-event (e.g., OS) and ordinal (e.g., RECIST) variables while Bayesian MCDA and SMAA require dichotomization of these outcomes. Even when the type I error rate is controlled, a constraint commonly requested by regulators, our simulation showed that MCADA outperforms existing MCDA methods. Although potentially counter-intuitive at first, our finding that SMAA has lower statistical power than either MCADA or probabilistic MCDA is in keeping with previous studies showing that SMAA accurately rejects high-risk and low-benefit treatments that other approaches may accept. Overall, these simulations indicate that SMAA is a more conservative MCDA approach. As a result, SMAA may be particularly well-suited to situations in which preferences vary between key stakeholders, which is not uncommon in clinical settings [Magliano et al., 2018, Stolker et al., 2014].

Our analyses of two case studies demonstrate that the proposed MCADA method can be valuable in decision-making, particularly when results from the primary outcome are inconclusive. In an early phase clinical trial comparing the safety and efficacy of LY2510924 and SOC combination therapy with SOC alone, the primary outcome of OS indicated a shorter duration in the treatment arm, though this difference lacked statistical significance. Relying solely on the trial’s primary outcome data did not provide a clear basis for deciding whether to continue or halt the trial. However, MCADA demonstrated that not only did the overall utility score favor the control arm, but the utility scores for each component consistently indicated a superior benefit-risk profile for the control arm compared to the treatment arm. The same trend was observed in the second case study.

The addition of the Emax and logistic transformation functions allows for non-linear modelling of the performance-utility relationship that is easily adjusted for each endpoint. Because these functions are specified for each variable, they have a straightforward interpretation. For example, clinicians may only be interested in achieving a certain level of a biomarker outcome, rather than maximizing it. MCADA allows specification of a utility function that aligns with clinical meaning. In contrast, other non-linear utility functions suffer from non-intuitive interpretation of the relationship between the treatment performances and the utility values [Menzies et al., 2022]. Our utilization of the Bayesian approach also facilitates the use of these non-linear utility functions. An appeal of using linear utility functions is that the total utility is a linear transformation of the criteria, so the statistical properties of the total utility are simpler to derive. This concern is addressed using a Bayesian approach, as modern sampling algorithms such as Markov chain Monte Carlo are available to characterize the distribution of non-linear total utilities.

The main strength of the MCADA methodology over existing benefit-risk assessment tools, such as MCDA [Saint-Hilary et al., 2017, 2018, Waddingham et al., 2016], is its ability to accommodate a wider range of data forms and use both linear and non-linear functions. By accommodating variables in their natural forms and uncertainty in estimates and allowing specification of each risk-benefit criterion’s utility function, MCADA can more accurately reflect what is known about a given clinical innovation and guide decision-making for a wider range of medical products, particularly in clinical settings [Chisholm et al., 2022, Hughes et al., 2007].

MCADA offers a comprehensive assessment approach for the efficient evaluation of medical products by employing a more holistic definition of success that considers treatment effect significance, clinical relevance, and favorable benefit-risk balance. There are limitations to the MCADA method. Due to its breadth and flexibility, MCADA is computationally intensive and as a result, the simulation study relied on a simulation of 2,500 trials. To address the limited number of simulations, we have reported Monte Carlo Standard Errors. Future work should consider ways to maintain the flexibility of MCADA while reducing computational requirements. We assumed a fixed utility weight for our MCADA analyses and did not consider other scenarios with unknown or uncertain utility weights. We also assumed utility to be linear across our simulation study analyses. As with other MCDA methods, MCADA treats all endpoints as additive and does not consider multiplicative interactions (i.e., that a given combination of outcomes may be more or less than the sum of its parts). Multiple sensitivity analyses need to be performed with varying utility values for different criteria. To address some of the limitations, MCADA may benefit from adding a Scalar-Loss Score (SLoS) utility function [Saint-Hilary et al., 2019]. Instead of using a linear utility score to find a treatment with the highest utility, the SLoS method aims to find a treatment with the lowest loss. In future studies, MCADA could be extended to allow for unknown and uncertain weights and interactions between criteria. Exploration of data in which efficacy is associated with higher toxicity, or other correlated endpoint scenarios, and the comparison of MCADA to other methods could be illustrative. Additionally, like Bayesian MCDA, MCADA assumes that risk-benefit criteria are uncorrelated [Waddingham et al., 2016]. This assumption may be unrealistic and future work should investigate its impact. One possible solution could be considering correlation among criterion using a sensitivity analysis, as in Bayesian MCDA. Lastly, we have assumed data to be non-missing in our simulations and missing at random in our analyses. As recommended by the editors, future work should investigate the impact of missing data on MCADA and other MCDA methods.

## Data Availability

This publication is based on research using information obtained from www.projectdatasphere.org, which is maintained by Project Data Sphere.

## Author contributions

JJHP had full access to all of the data in the study and takes responsibility for the integrity of the data and the accuracy of the data analysis.

Concept and design: JJHP

Acquisition, analysis, or interpretation of data: QV, RKM, OH, EJM, and JJHP

Drafting of the manuscript: QV, RKM, OH, EJM, and JJHP

Critical revision of the manuscript for important intellectual content: QV, RKM, OH, EJM, and JJHP

Statistical analysis: QV, OH, and JJHP

Software development: OH, QV and JJHP

Obtained funding: JJHP

Administrative, technical, or material support: OH, JJHP

Supervision: JJHP

## 7 Acknowledgements

We thank Dr. Awa Diop for providing their insight in data generating mechanisms for simulating time-to-event analyses. We thank Dr. Vinusha Kalatharan for their critical review of a previous version of the manuscript. We also thank the associate editors for their constructive feedback.

